# Interim influenza vaccine effectiveness estimates during the 2023 Southern Hemisphere season

**DOI:** 10.1101/2023.09.07.23295154

**Authors:** J Gonzalo Acevedo-Rodriguez, Carlos Zamudio, Noah Kojima, Fiorella Krapp, Pablo Tsukayama, C Stefany Neciosup-Vera, Eduardo Gotuzzo

## Abstract

Our findings suggest that a substantial number of people sought care for CLI during an early Southern Hemisphere season during which A/H1N1pdm09 clade 6B.1A.5a.2a viruses predominated. The minority vaccinated benefitted from the A/Victoria/2570/2019 (H1N1)pdm09-like antigen which afforded good protection against illness.

## Main Text

Evaluations of vaccine effectiveness (VE) against influenza in the Southern Hemisphere (SH) can assist that region of the world and provide insights into the anticipated performance of Northern Hemisphere (NH) vaccines.^1,2^ Using population-based surveillance data for COVID-19, we estimated VE against medically attended influenza in Lima, Peru.

From January 1 to July 31, 2023, our population-based surveillance identified patients seeking care at two primary health centers in San Juan de Lurigancho, Lima. Patients presenting with COVID-19-like illness (CLI, defined as onset ≤7 days of ≥2 signs or symptoms, e.g., documented or reported fever, chills, rigors, myalgia, headache, and sore throat) were asked for written consent to participate. Enrolled participants reported preexisting condition and influenza vaccine histories and provided nasopharyngeal swabs for influenza virus and SARS-CoV-2 testing through multiplex qPCR.^3^ Whole-genome sequencing was conducted for positive specimens with a cycle threshold value ≤25 using the Illumina Respiratory Virus Panel.

Incidence of medically attended influenza illness was estimated by dividing the number of patients with CLI that consented to participate by the population assigned to the health centers (i.e., 91,801) and adjusting for enrollment (supplement). Patients who self-reported influenza vaccination from May 2022 until >14 days before enrollment were classified as vaccinated. VE against medically attended influenza was calculated using a test-negative design.^4^ Cases were defined as patients with a test-positive for influenza by qPCR test and controls as patients test-negative for both influenza and SARS-CoV-2. The study was approved by the institutional review board of Universidad Peruana Cayetano (PRISA repository: EI0002439) Centers for Disease Control and Prevention non-research determination 0900f3eb81ecbe9f.

During study period, 923 (13.9%) of 6,660 persons who sought outpatient care met criteria for CLI, and 567 (61.4%) were enrolled and had testing for influenza viruses and SARS-COV-2 (Table 1). Influenza was detected in 106 (18.7%) as early as January, months before the historical May start of influenza seasons in Lima (Figure 1). Seventy-nine (75%) patients with CLI tested-positive for influenza A and 27 (25%) for B. We sequenced 32 influenza A specimens; 29 (90.6%) were A(H1N1)pdm09 clade 6B.1A.5a.2a (Table 1). The cumulative incidence of medically attended influenza per 1,000 population was 10.1 (95% confidence interval [95%CI]: 8.2–12.0) for influenza A and 3.5 (95%CI: 2.3–4.6) for influenza B. Only 112 (19.7%) patients were vaccinated against influenza. The age and month adjusted VE against medically attended influenza was 65.1% (95%CI: 26.5–83.4); 58.2% (95%CI: 5.1–81.6) for influenza A and 77.6% (95%CI: 0–95.5) for B.

**Table 1.**
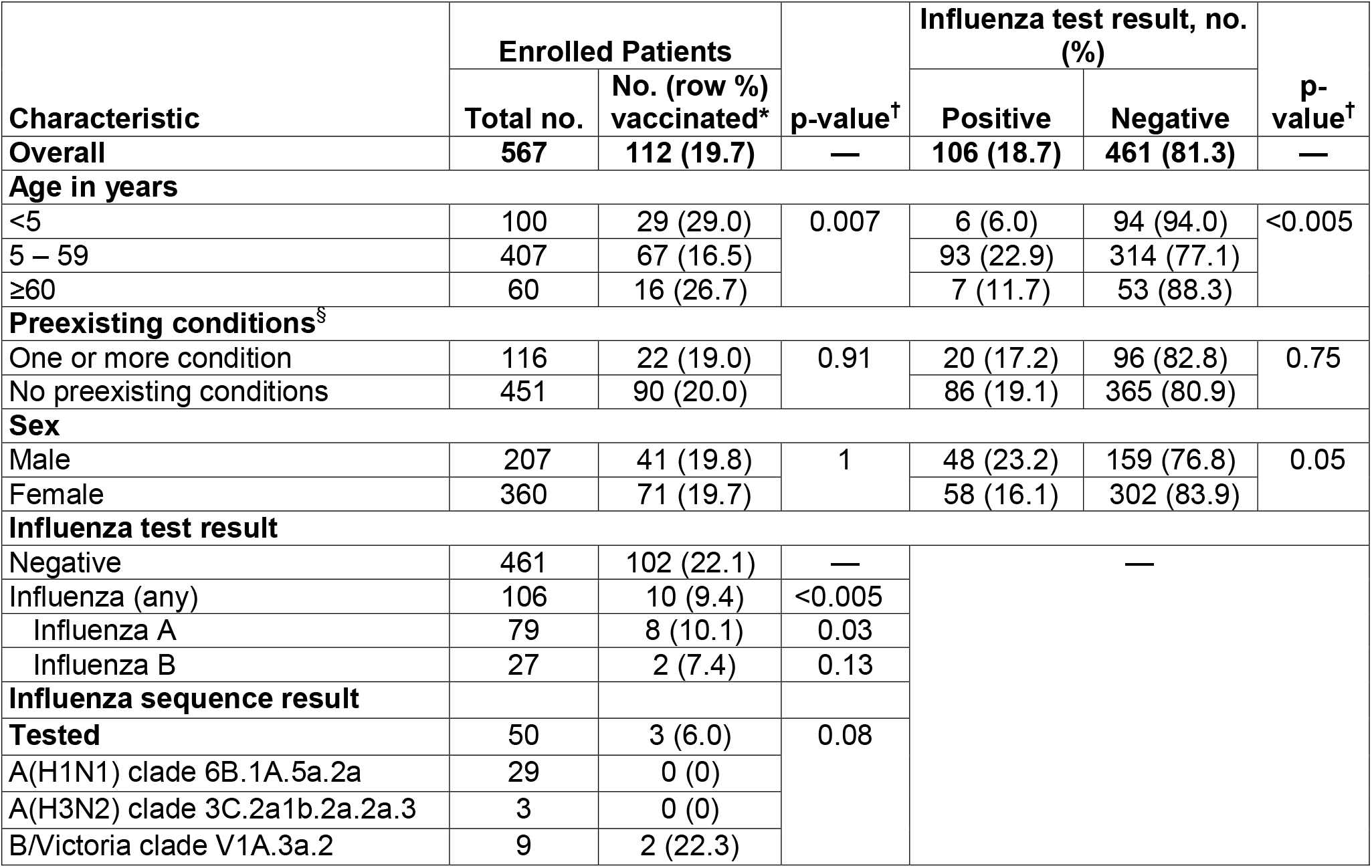
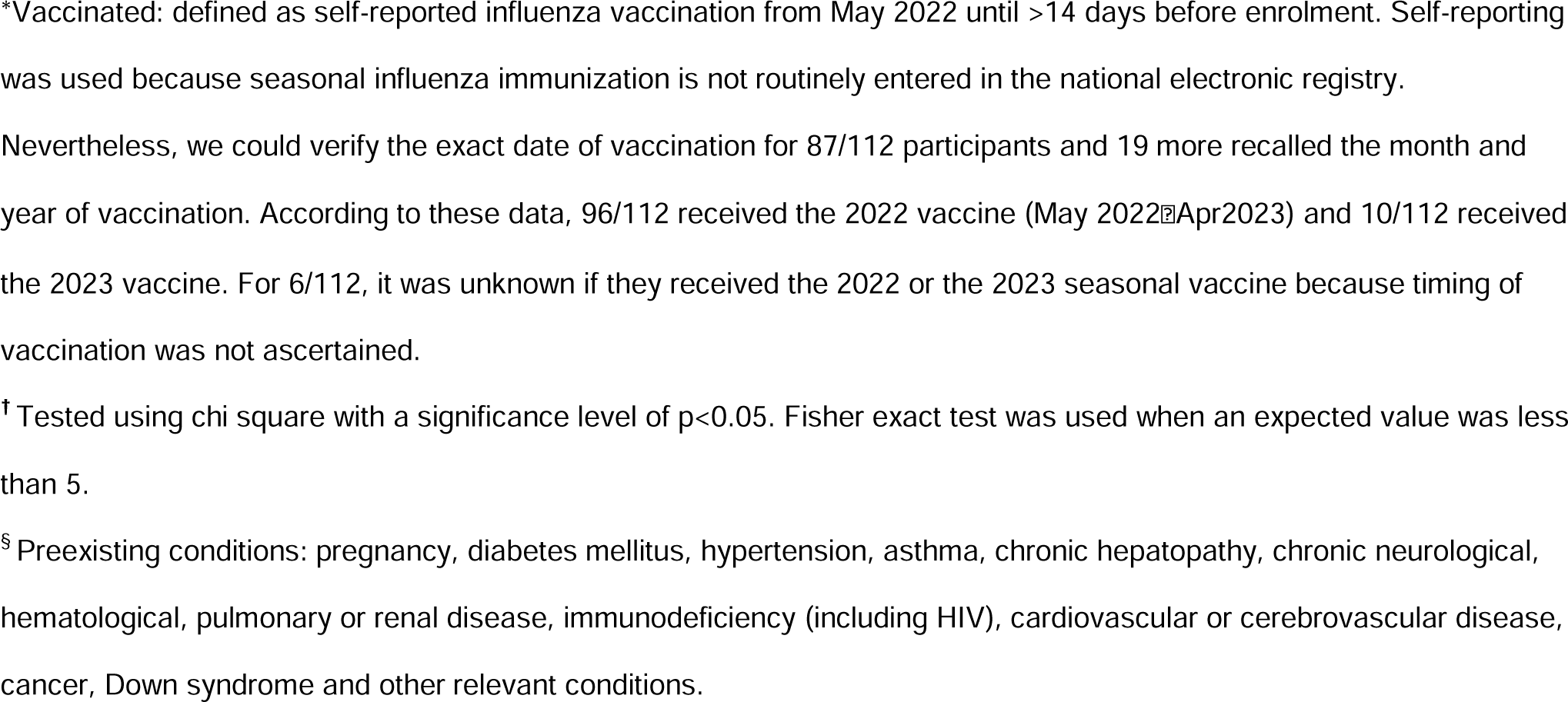
Influenza test results and seasonal vaccination status among patients presenting in two primary care centers with COVID-19-like illness in Lima, Peru, by selected characteristics (N = 567), January–May 2023.

**Figure 1.**
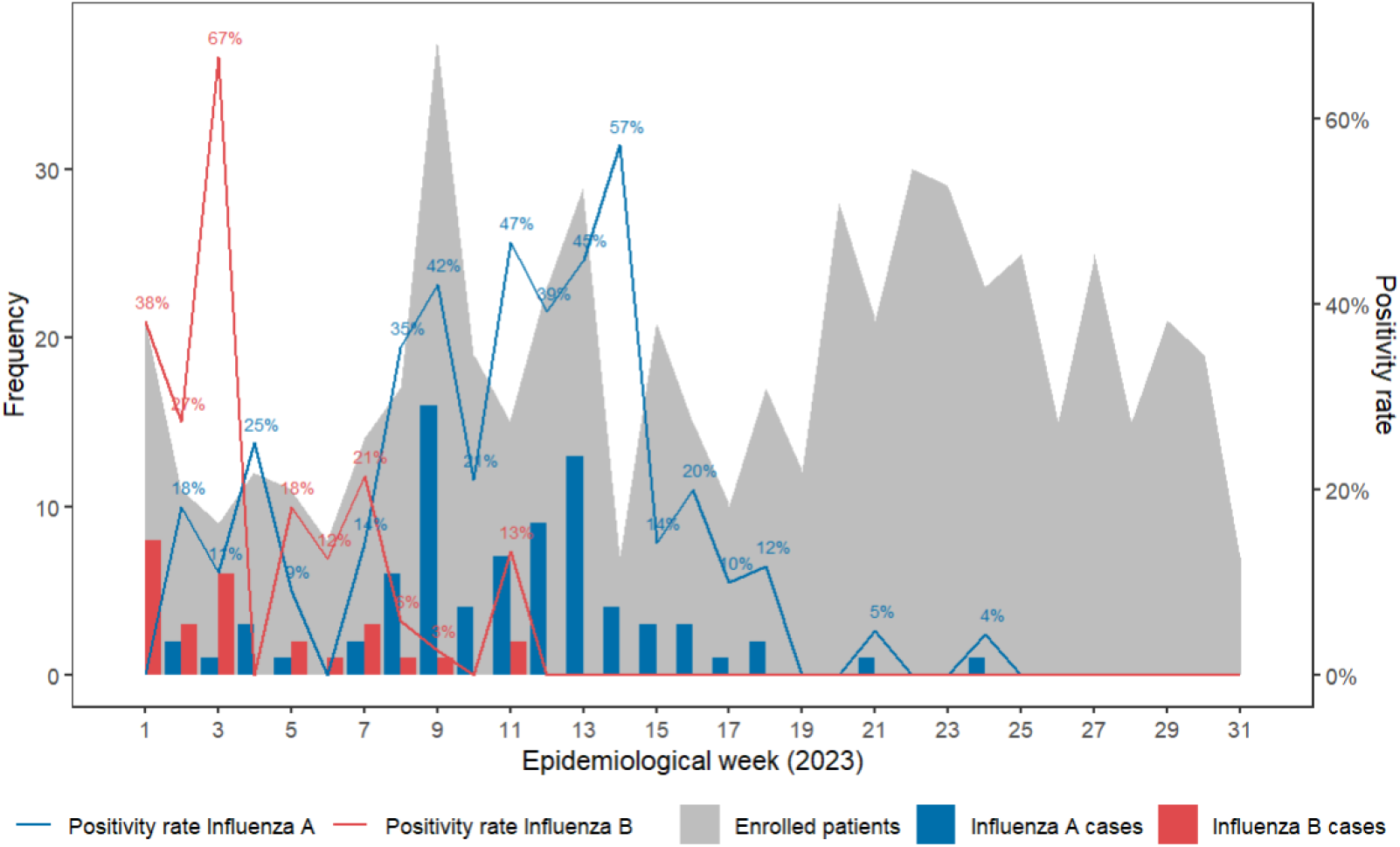
Weekly laboratory-confirmed incident influenza cases and positivity rate at two primary care health centers from patients who presented with COVID-19-like illness in Lima, Peru, January–May 2023

During the 2023 SH season, a substantial number of people sought care for influenza, but incidence was lower than reported among persons seeking care for influenza-like illness from before COVID-19.^5^ 2023 SH VE against influenza was similar to that of the 2022 SH season^1,2^ and the 2022-2023 U.S. season^6^ even though A(H1N1)pdm09 clade 6B.1A.5a.2a viruses predominated in 2023 whereas influenza A(H3N2) clade 2.a3 viruses predominated in 2022.^7^ The A/Victoria/2570/2019 (H1N1)pdm09-like virus in the SH vaccine conferred a 65% risk reduction in medically attended influenza, yet fewer than one in five persons in this evaluation reported receiving an influenza vaccine during 2023.

Our findings are subject to limitations. First, in the absence of healthcare utilization surveys, it is unclear if the population assigned to our clinics always sought care there versus other clinics. Second, only half of specimens had cycle threshold value ≤25; thus, it is possible that we missed clades. Finally, confounding might remain in our analysis despite our statistical models.

In conclusion, our findings suggest that mostly unvaccinated people sought care early during SH season for illnesses predominantly attributable to A/H1N1pdm09 clade 6B.1A.5a.2a viruses. Persons vaccinated benefitted from the A/Victoria/2570/2019 (H1N1)pdm09-like antigen, which afforded good protection against illness. If the same viruses predominate in the NH, our estimates of 2023 SH influenza vaccine effectiveness suggest that 2023-2024 NH influenza vaccines, which include a similar strain composition, could provide similar protection during the upcoming influenza season.

## Data Availability

All data produced in the present study are available upon reasonable request to the authors.

## Declarations

None.

## Disclaimer

The findings and conclusions in this report are those of the authors and do not necessarily represent the official position of the Centers for Disease Control and Prevention.

## Author Contribution

CZ and EG had full access to all the data in the study and take responsibility for the integrity of the data and the accuracy of the data analysis. Data will be shared by contacting the corresponding author upon request for research purposes. JGA-R, CZ, FK, FM, PT and EG conceived and designed the study, EM coordinated laboratory analysis, PT performed genomic sequencing, PA, FG and RC coordinated data collection and management, CSN-V and GSR performed the statistical analysis. All authors wrote and approved the final manuscript. NK and EGL helped draft, edit and review the manuscript. LC, AF and EAB helped interpret findings and edit the manuscript.

## Acknowledgements

None

