## Supplemental for "Interim influenza vaccine effectiveness estimates during the 2023 Southern Hemisphere season"

**Supplemental Materials**

**Estimation of Incidence**

The point estimation of the incidence was calculated using the following formula:

$$\frac{n_{p} + n_{u} \hat{p}}{N}=\frac{n_{p}+n_{u}\frac{n_{p}}{n}}{N}$$

The variance of the estimator is approximated by the sum of: a) the variance of the number of observed cases and the variance of the un (approximately) observed cases:

$$\hat{Var}\left( \frac{n_{p} + n_{u} \frac{n_{p}}{n}}{N} \right)= \frac{\left( 1+\frac{n_{u}}{n} \right)^{2}n^{2}}{N^{2}}\frac{\hat{p}(1-\hat{p})}{n}$$

An interval estimation is given by:

$$\frac{n_{p}+n_{u}\frac{n_{p}}{n}}{N} \mp1.96\sqrt{\frac{\left( 1+\frac{n_{u}}{n} \right)^{2}n^{2}}{N^{2}}\frac{\hat{p}(1-\hat{p})}{n}}$$

Number of enrolled patients: $n$

Number of positive cases among enrolled patients: $n_{p}$

Proportion of positive cases among the enrolled cases: $\hat{p}$

Number of non-enrolled CLI cases: $n_{u}$

The number of non-enrolled positives cases is estimated by $n_{u}$ $\hat{p}$

N: population assigned by Regional Health Direction of the Ministry of Health

to both centers is 91,801.

*Note:* San Juan de Lurigancho is a district with 1,196,099 inhabitants. There are 36 public primary care centers in this district, each health center is assigned a specific catchment area that is mutually exclusive. The population living within the catchment area -per the last national census- is the population assigned to each center by Regional Health Direction of the Ministry of Health.

**Supplemental Figure 1.** Influenza subtypes, lineages and clades determined by whole genome sequencing from positive respiratory samples collected at two primary care health centers from patients who presented with COVID-19-like illness in Lima, Peru (epidemiological weeks 1–30, 2023)

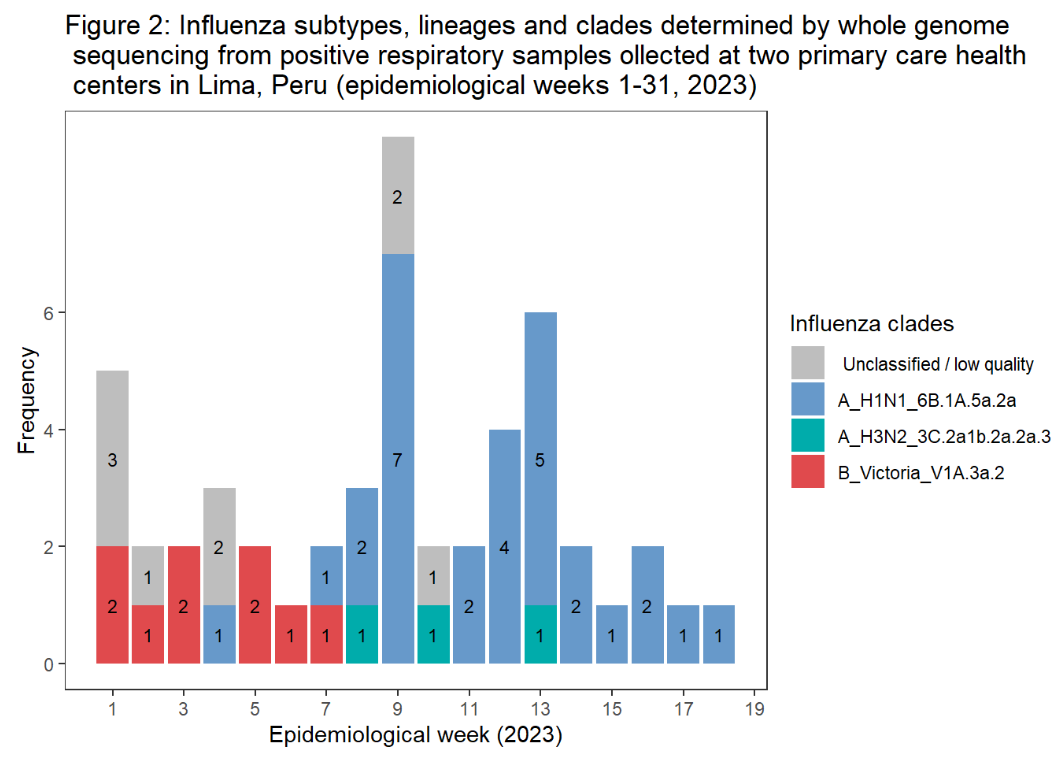
